# Polypharmacy and mortality in older persons: findings from a sub-cohort of SABE Colombia

**DOI:** 10.64898/2026.08.19.26360848

**Authors:** Hernán-David García, Carolina Giraldo Benítez, Jorge Hernando Donado, Pedro Hernández, Camila Vélez, Luis-Ángel Toro, Carmen-Lucía Curcio

## Abstract

**Background:** Polypharmacy is an escalating global health challenge, yet longitudinal evidence regarding its impact on mortality in Latin American aging populations remains limited. This study evaluated the association between medication burden and all-cause mortality among community-dwelling older adults in a rapidly aging region of Colombia.

**Methods:** A longitudinal analysis was conducted using a sub-cohort of 4,110 participants (aged ≥60 years) from the SABE Colombia survey (Antioquia, Caldas, Risaralda, and Quindío). Vital status was adjudicated via the National Health System Resources Administrator (ADRES) database over a mean follow-up of 79 months. Polypharmacy was defined as the concurrent use of 5–9 medications and excessive polypharmacy as ≥10. Extended Cox proportional hazards models were employed to estimate hazard ratios (HR), adjusting for sociodemographic factors, multimorbidity, and functional dependency.

**Results:** At baseline, 20.2% of participants presented polypharmacy and 2.1% excessive polypharmacy. A total of 1,092 deaths (26.6%) were recorded during follow-up. After multivariable adjustment, both moderate polypharmacy (HR 1.17; 95% CI 1.02–1.31; p=0.029) and excessive polypharmacy (HR 1.82; 95% CI 1.34– 2.47; p<0.001) were identified as independent predictors of mortality. Notably, the risk was markedly higher at the ≥10 medication threshold, suggesting a non-linear relationship between pharmacological burden and survival.

**Conclusions:** Polypharmacy is a significant and independent predictor of mortality in Colombian older adults, with the risk nearly doubling in cases of excessive medication use. These findings underscore the urgent need for structured medication review and deprescribing interventions tailored to resource-constrained healthcare systems to mitigate the risks associated with high pharmacological accumulation.

## Introduction

Aging is frequently associated with the onset of multimorbidity, which leads to the use of multiple medications by older patients. This consequently results in polypharmacy, defined as the simultaneous use of multiple drugs [1]. The literature lacks a universally accepted definition of polypharmacy. Approximately 143 different definitions have been identified, the vast majority of which are purely numerical. This reliance on the count of medications consumed per individual introduces considerable heterogeneity across studies. Polypharmacy thresholds range from the consumption of two or more medications up to eleven or more medications, with similarly heterogeneous classifications. Other numerical definitions also incorporate the duration of treatment or the total number of medications consumed during a hospital stay [2].

Beyond numerical counts, non-numerical definitions focus on clinical context. These include necessary polypharmacy, which justifies additional medications for optimized management, and qualitative polypharmacy, defined by the inclusion of potentially inappropriate medications. More specialized terms include psychotropic polypharmacy (the use of two or more psychotropic agents) and the distinction between appropriate and inappropriate polypharmacy, based on the clinical necessity and indication of the regimen [3]. Despite this conceptual breadth, the most widely accepted clinical standard defines polypharmacy as the simultaneous use of five or more medications, with ‘excessive polypharmacy’ reserved for regimens of ten or more [4,5].

Polypharmacy must not be intrinsically equated with suboptimal or inappropriate prescribing. In the context of multimorbidity, complex therapeutic regimens are often clinically justified and align with evidence-based guidelines for managing co-occurring chronic conditions. Nevertheless, an escalating medication burden inherently increases treatment complexity, elevating the risk of drug-drug interactions and complicating both patient adherence and clinical management. These challenges persist even when individual prescriptions are deemed appropriate, suggesting that the cumulative effect of multiple medications transcends the safety profile of its isolated components [2].

Global estimates of polypharmacy prevalence exhibit significant variability, typically ranging from 30% to 60% within the geriatric population [6]. Regional data from North America indicate that 22.4% of individuals in the United States and 18.8% in Canada aged 40–79 years utilize five or more medications concurrently [7]. Within this international landscape, Colombia presents a distinct pharmacological profile; population-based evidence suggests a polypharmacy prevalence of 31% (5–9 medications), while excessive polypharmacy (≥10 medications) affects 1.8% of older adults [8]. These figures underscore the substantial medication burden in the Colombian context and emphasize the urgent need to evaluate its long-term clinical consequences, particularly in terms of survival.

The clinical implications of polypharmacy are multifaceted, encompassing a heightened susceptibility to adverse drug reactions, complex pharmacological interactions, and suboptimal treatment adherence. Beyond these immediate risks, an elevated medication burden serves as a critical driver for geriatric syndromes, including falls, frailty, and the impairment of objective physical performance— specifically gait speed, chair-rise capability, and grip strength [9–11]. These associations are robustly supported by international longitudinal evidence: while Australian cohorts have linked polypharmacy to an increased incidence of falls and fractures [12], studies in Japan and the United States have further established its role as a significant predictor of cognitive decline, hospitalization, and all-cause mortality [13–15].

Despite the documented risks of polypharmacy in high-income settings, its clinical impact may diverge in middle-income countries like Colombia, where rapid demographic aging and fragmented healthcare delivery create unique challenges for medication safety and deprescribing. In such resource-constrained environments, polypharmacy often reflects a complex interplay between clinical multimorbidity and systemic barriers to continuity of care. Consequently, understanding its association with mortality is essential for developing targeted, locally-adapted interventions. To address this critical evidence gap, this study utilizes longitudinal data from the SABE Colombia survey to evaluate the relationship between polypharmacy and all-cause mortality among community-dwelling older adults.

## Methods

### Population

Data for this study were obtained from the Colombian Survey on Health, Well-Being, and Aging (SABE Colombia), a cross-sectional survey conducted between 2014 and 2015. The survey was developed by the Ministry of Health and Social Protection and the Administrative Department of Science, Technology, and Innovation (COLCIENCIAS), in collaboration with Universidad del Valle and Universidad de Caldas. Its primary objective was to characterize the health status of the older population in Colombia and to generate indicators related to aging.

The target population for this study comprised individuals aged 60 years and older residing in Colombia who participated in the SABE Colombia survey, a nationally representative study (23694 participants). For the purposes of this analysis, a subsample (N = 4110) was selected, focusing on four departments: Antioquia, Caldas, Risaralda, and Quindío. This subsample was chosen based on the following criteria: (i) these departments exhibit the highest aging indices in the country; (ii) they share similar sociodemographic, cultural, and economic characteristics; and (iii) this subgroup had more complete data regarding the mortality outcome [16]. The methodological document of the National Health, Well-being, and Aging Survey SABE Colombia can be consulted at the following <u>link</u>.

### Variables

The primary dependent variable was all-cause mortality, determined by verifying the vital status of each participant through the official registry of the Administrator of Resources of the General Social Security System in Health (ADRES). It is important to note that this database comprehensively covers individuals affiliated with the contributory and subsidized health regimes, while excluding those enrolled in ‘special regimes.’ Survival time, or time-to-event, was defined as the number of months elapsed between the baseline interview and the date of death. The study achieved a robust mean follow-up period of 79 months, reaching a maximum of 94 months.

To account for potential confounding, the analysis integrated a wide array of covariates. Sociodemographic factors included age (measured in completed years), sex, interview date, and department of residence. Socioeconomic status was operationalized through Colombia’s six-tier stratification system, where tiers 1 (very low) to 6 (high) serve as a proxy for household conditions, public service availability, and urban-environmental characteristics. Additionally, educational attainment was recorded as total years of completed schooling, and housing conditions were dichotomized into urban or rural settings.

Health-related and functional measures were meticulously assessed. The burden of chronic diseases was derived from self-reported medical diagnoses. Functional status was evaluated using two validated instruments: the Barthel Index, which assesses physical dependency through Basic Activities of Daily Living (B-ADL) such as bathing, dressing, and feeding; and the Lawton and Brody Scale, which captures Instrumental Activities of Daily Living (I-ADL), including more complex tasks like medication management, financial handling, and transportation use. Participants were classified as functionally dependent if they reported difficulty or inability in at least one activity across either scale.

Regarding medication data, exposure was determined by direct verification of medical prescriptions or medication packaging; in cases where physical evidence was unavailable, data were obtained through participant self-report. Medications were categorized by therapeutic class, including neurological, cardiovascular (covering hypertension, diabetes, and dyslipidemia), respiratory, gastrointestinal, and analgesics, among others. Polypharmacy was defined as the concurrent use of five or more medications, while excessive polypharmacy was reserved for the use of ten or more, both calculated as person-level daily counts at baseline to ensure a longitudinal perspective of the pharmacological burden.

## Statistical Analysis

### Descriptive and univariate analysis

A descriptive analysis was conducted for all study variables. Absolute and relative frequencies were reported for qualitative variables, while measures of central tendency (mean and standard deviation) were used for quantitative variables. The primary exposure variable, polypharmacy, was introduced into the survival models as a categorical variable with three levels: 1) 0−4 medications (Reference group); 2) 5−9 medications (Polypharmacy); and 3) ≥10 medications (Excessive Polypharmacy). The Kaplan–Meier method was used to analyze mortality across these three groups, followed by the Log-Rank test to assess crude survival differences.

### Handling of complex survey design and weights

We conducted a secondary analysis using a sub-cohort of the SABE Colombia survey, restricted to four departments (Antioquia, Caldas, Risaralda, and Quindío) due to data completeness for mortality follow-up. We acknowledge that the original SABE Colombia survey utilized a complex, multi-stage sampling design. However, we did not apply the original survey weights in the Cox proportional hazards models. This decision was based on the fact that our primary objective was to estimate the association (internal validity) between polypharmacy and mortality within the observed cohort, rather than to produce population-level estimates of prevalence or weighted hazard ratios. Consequently, the effect estimates (Hazard Ratios) pertain specifically to this cohort and are not nationally representative.

### Covariate selection and model specification

To estimate the relative risk of death, we applied a multivariable Cox regression model, using polypharmacy as the primary variable of interest. The model was constructed by adjusting for sociodemographic (age) and clinical/functional variables (multimorbidity and functional dependence -using the Barthel/B-ADL scale-).

Variables such as socioeconomic status, education level, housing characteristics, and Instrumental Activities of Daily Living (I-ADL) were tested but ultimately excluded from the final adjusted model. This decision prioritized model parsimony, avoided potential multicollinearity (particularly between B-ADL and I-ADL), and was supported by the finding that their inclusion did not significantly alter the effect estimates for polypharmacy, suggesting that their confounding effect was largely captured by the inclusion of multimorbidity and basic functional dependence.

Stratified analyses by specific comorbidity types were not performed, as the study focused on overall medication burden at the individual level. Moreover, stratification by individual diseases would have resulted in small subgroup sizes and reduced statistical power, particularly for excessive polypharmacy, and may have led to collinearity given the high coexistence of chronic conditions in older adults.

### Extended Cox model for proportional hazards correction

The proportional hazards (PH) assumption, a core requirement for Cox regression, was formally assessed using Schoenfeld residuals (via the cox.zph function in R). Significant violations of this assumption were detected for the global model and specifically for the covariates B-ADL dependency and multimorbidity (p<0.001).

To address these violations and ensure valid estimation of the effects, an Extended Cox Proportional Hazards Model was fitted. Non-proportionality for B-ADL dependency and multimorbidity was modeled by including time-dependent interaction terms (covariate⋅log(time+1)). This allowed for time-varying hazard ratios for these specific factors while providing stable and accurate estimates for the effect of polypharmacy. Statistical significance was set at a two-tailed p-value <0.05. Statistical analyses were conducted using Jamovi software (Version 2.6) and R.

## Results

### Cohort characteristics

We analyzed 4110 older adults (mean age 70.7 ± 8.2 years), of whom 61.2% were women. Most participants lived in urban areas (83.1%) and belonged to the lowest socioeconomic stratum (70.9%). Nearly one in five were dependent on basic activities of daily living (B-ADL; 19.1%), and over one third were dependent on instrumental ADL (37.4%). Multimorbidity (≥ 2 chronic conditions) was present in 44.4%. During follow-up, 1018 participants died (24.7%).

### Medication exposure

Polypharmacy (≥ 5 medications) was documented in 20.2% of participants; 2.1% used ≥ 10 medications. Cardiovascular agents were the most frequently reported class (61.9%), followed by analgesics (18.2%) and gastrointestinal medications (17.6%). Neurological and pulmonary medications were reported by 9.0% and 5.9%, respectively; 3.7% reported over-the-counter products. “Other” medications were recorded for 27.4% of the cohort. Baseline characteristics of the study population are shown in Table 1.

**Table 1.**
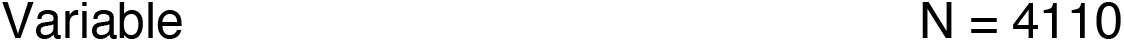
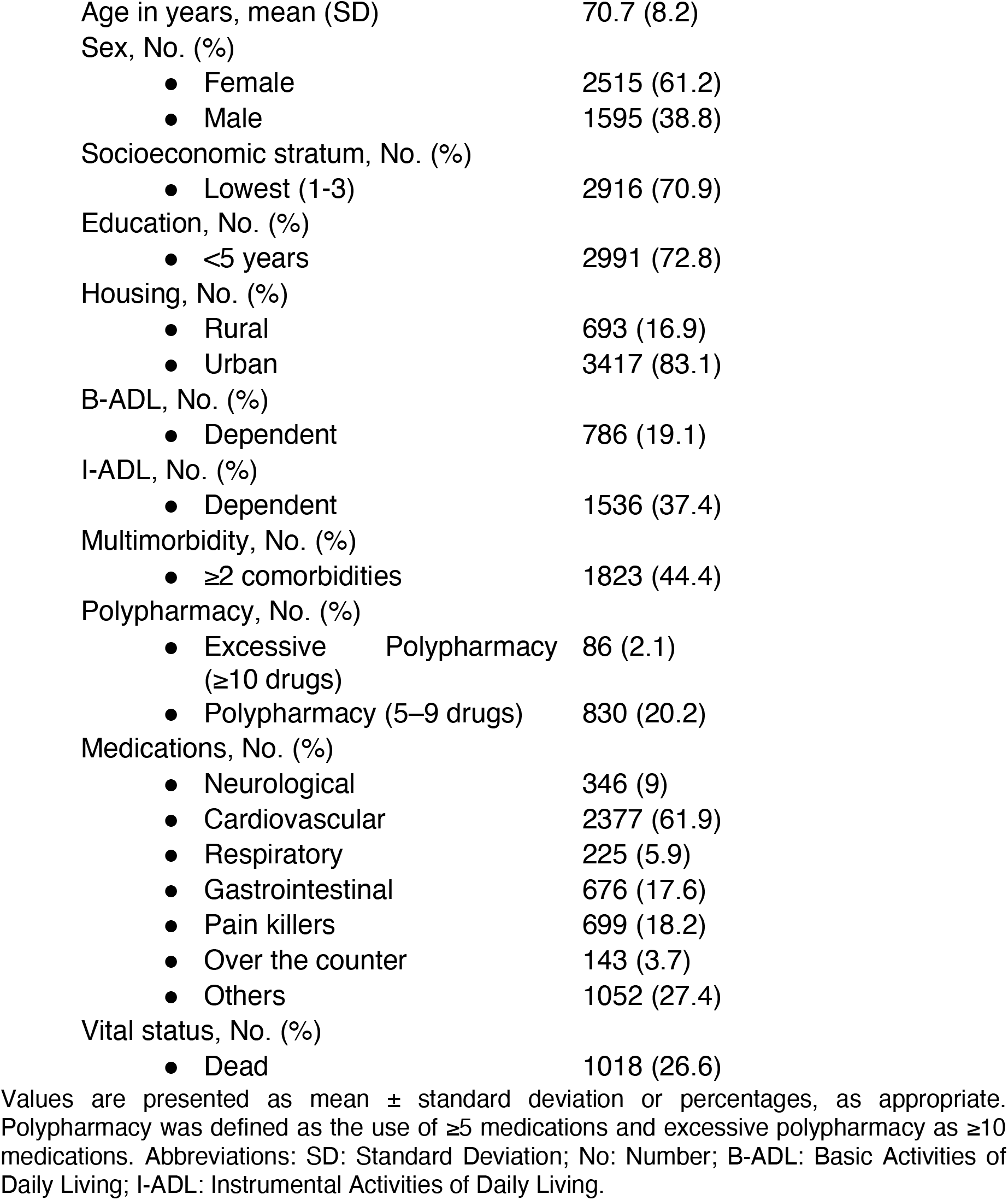
Baseline characteristics of community-dwelling older adults included in this cohort (N = 4110)

|  |  |
| --- | --- |
| Age in years, mean (SD) | 70.7 (8.2) |
| Sex, No. (%) |  |
| • Female | 2515 (61.2) |
| • Male | 1595 (38.8) |
| Socioeconomic stratum, No. (%) |  |
| • Lowest (1-3) | 2916 (70.9) |
| Education, No. (%) |  |
| • <5 years | 2991 (72.8) |
| Housing, No. (%) |  |
| • Rural | 693 (16.9) |
| • Urban | 3417 (83.1) |
| B-ADL, No. (%) |  |
| • Dependent | 786 (19.1) |
| I-ADL, No. (%) |  |
| • Dependent | 1536 (37.4) |
| Multimorbidity, No. (%) |  |
| • $\geq 2$ comorbidities | 1823 (44.4) |
| Polypharmacy, No. (%) |  |
| • Excessive Polypharmacy ($\geq 10$ drugs) | 86 (2.1) |
| • Polypharmacy (5–9 drugs) | 830 (20.2) |
| Medications, No. (%) |  |
| • Neurological | 346 (9) |
| • Cardiovascular | 2377 (61.9) |
| • Respiratory | 225 (5.9) |
| • Gastrointestinal | 676 (17.6) |
| • Pain killers | 699 (18.2) |
| • Over the counter | 143 (3.7) |
| • Others | 1052 (27.4) |
| Vital status, No. (%) |  |
| • Dead | 1018 (26.6) |

Values are presented as mean ± standard deviation or percentages, as appropriate. Polypharmacy was defined as the use of $\geq 5$ medications and excessive polypharmacy as $\geq 10$ medications. Abbreviations: SD: Standard Deviation; No: Number; B-ADL: Basic Activities of Daily Living; I-ADL: Instrumental Activities of Daily Living.

### Survival analyses

Kaplan-Meier survival curves, stratified by the three polypharmacy categories (0–4, 5–9, and ≥10 medications), showed a statistically significant difference in survival probability among the groups throughout the 79-month follow-up period (Log-Rank Test, p<0.0001). The survival probability was highest for participants using 0−4 medications and lowest for those with excessive polypharmacy (≥10 medications), demonstrating a clear dose-response effect of medication burden on mortality risk (See Figure 1).

**Figure 1.**
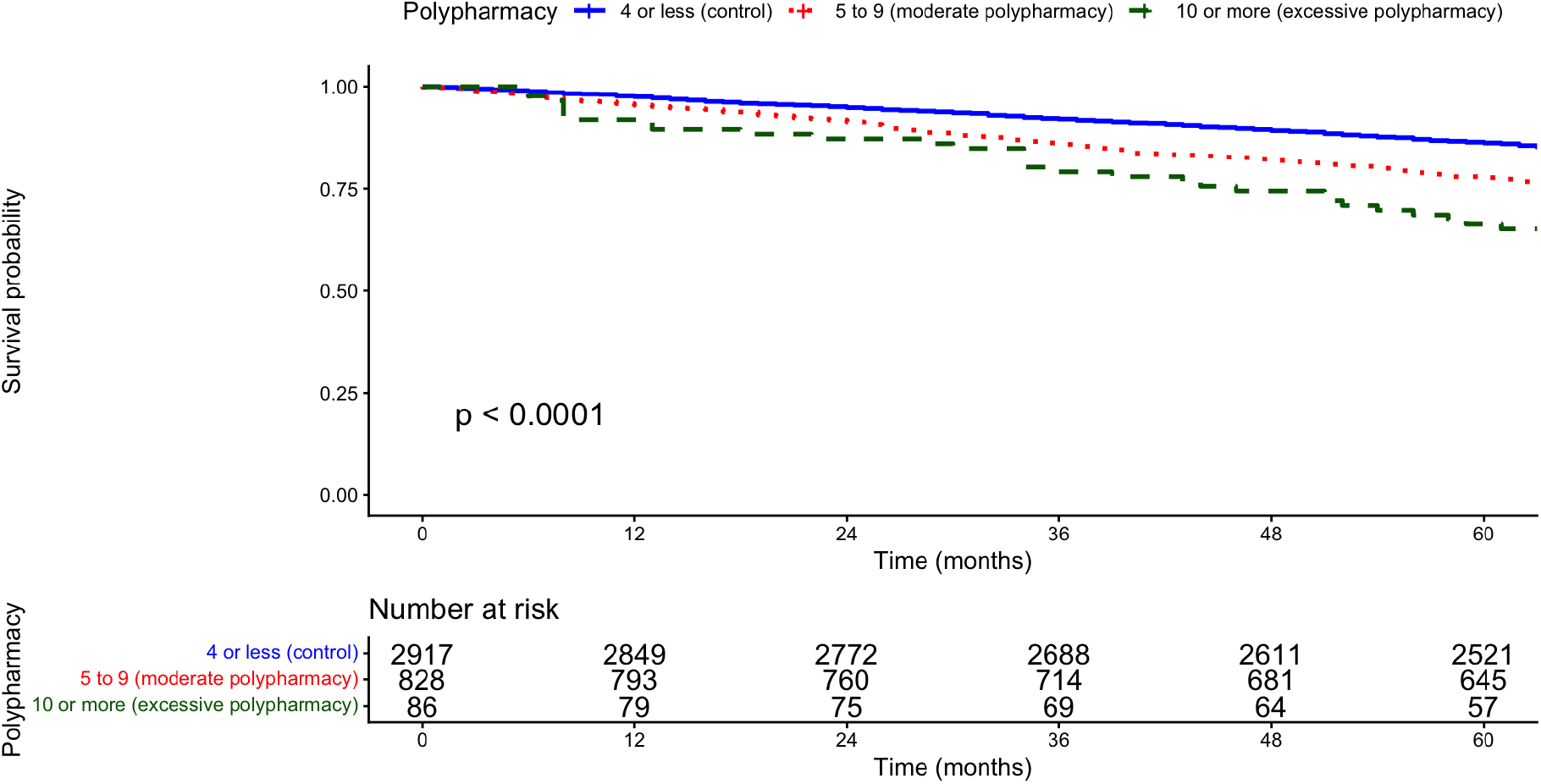
Kaplan-Meier Survival Curves stratified by three levels of polypharmacy exposure.

In the univariable Cox models, polypharmacy (≥5 medications) was associated with a 68% higher risk of mortality (HR 1.68, 95% CI 1.47–1.91; p < 0.001). By contrast, excessive polypharmacy (≥10 medications) conferred an even greater risk, more than doubling the hazard of death (HR 2.56, 95% CI 1.91–3.43; p < 0.001). Given the violation of the proportional hazards assumption for BADL dependency and multimorbidity, results are presented from an extended Cox model incorporating time-dependent effects. To assess potential multicollinearity among covariates in the multivariable model, a collinearity diagnostic was performed. All variables demonstrated a Variance Inflation Factor (VIF) below the commonly accepted threshold of 10, indicating no significant multicollinearity and supporting the stability and interpretability of the regression estimates.

After adjusting for age, B-ADL dependency, and multimorbidity and modeling the non-proportional hazards for the latter two via time-dependent terms moderate polypharmacy (≥5 medications) was associated with a statistically significant increase in mortality risk (HR 1.17; 95% CI: 1.02–1.35; p = 0.029). In contrast, excessive polypharmacy (≥10 medications) was associated with a substantially higher mortality risk (HR 1.82; 95% CI: 1.35–2.45; p < 0.001). Functional dependence and multimorbidity exhibited strong early associations with mortality, with a progressive attenuation of their effects over time. The association between polypharmacy and mortality is presented in Table 2.

**Table 2.** Association between polypharmacy and all-cause mortality in older adults from the cohort.

| Variable | Crude HR<br>(95% IC) | Adjusted HR | 95% CI | P-value |
| --- | --- | --- | --- | --- |
| 0–4 Medications (Reference) | 1 | 1 | — | — |
| 5–9 Medications<br>(Polypharmacy) | 1.68 (1.47-1.91) | 1.17 | 1.02 – 1.35 | 0.029 |
| ≥ 10 Medications (Excessive<br>Polypharmacy) | 2.56 (1.91-3.43) | 1.82 | 1.35 – 2.45 | < 0.001 |
| Age (per year increase) |  | 1.08 | 1.07 – 1.09 | < 0.001 |
| Multimorbidity (≥ 2 diseases) |  | 2.79 | 1.51 – 5.15 | 0.002 |
| B-ADL Dependency |  | 3.36 | 1.85 – 6.09 | < 0.001 |
| Time-dependent<br>multimorbidity effect |  | 0.81 | 0.68 – 0.95 | 0.009 |
| Time-dependent B-ADL effect |  | 0.82 | 0.70 – 0.97 | 0.017 |

Hazard ratios (HR) and 95% confidence intervals (CI) were estimated using Extended Cox proportional hazards model, adjusted for age, multimorbidity, and functional status (B-ADL).. Time-dependent covariates were modeled using the functional form $x \cdot \log(t + 1)$ , where $x$ represents the baseline value of the variable of interest and $t$ denotes time since baseline.

## Discussion

These findings confirm a graded association between medication burden and mortality, which should be interpreted within both clinical and health-system contexts. This study provides evidence on the relationship between polypharmacy and mortality among older adults in four Colombian departments. Its contribution does not lie in merely reaffirming the association between polypharmacy and mortality, which has been well documented, but rather in contextualizing this relationship within a middle-income country and delineating clinically meaningful gradients of risk. Our results demonstrate that although polypharmacy (≥5 medications) is associated with increased mortality, excessive polypharmacy (≥10 medications) confers a substantially higher risk, even after adjustment for multimorbidity and functional status.

Our results are consistent with previous studies. In Brazil, the SABE survey showed that polypharmacy was associated with reduced five-year survival [17]. Internationally, evidence from Europe and North America has demonstrated that polypharmacy prevalence often exceeds 40% and is linked to adverse outcomes such as falls, frailty, hospitalization, and mortality [18-20]. A Colombian study reported that 31% of older adults experienced polypharmacy and 1.8% excessive polypharmacy [8]. Taken together, these findings reinforce that while moderate polypharmacy signals vulnerability, excessive polypharmacy is particularly dangerous and should be a priority target for intervention.

From a clinical perspective, these findings highlight the need to prioritize systematic medication review as a core component of care for older persons, particularly in primary care and geriatric services. The observed association between higher medication burden and mortality supports the implementation of structured interventions, including the use of explicit prescribing tools to optimize medication appropriateness. Deprescribing strategies and shared decision-making can help reduce inappropriate medication use, improve adherence, and prevent drug–drug interactions. Incorporating pharmacists into primary care teams has shown promise in identifying potentially inappropriate prescriptions and supporting safer prescribing practices [15]. Furthermore, decision-support tools integrated into electronic health records may help clinicians balance therapeutic benefits with risks in complex older patients.

The significant effect of polypharmacy (≥5 medications) suggests that interventions should not focus solely on those with extreme medication counts but also on patients at earlier stages, where preventive deprescribing may avert progression to excessive polypharmacy. Clinicians must pay special attention to high-risk groups, including frail individuals, those with multiple chronic conditions, and those with dependence in daily activities, in whom therapeutic inertia must be avoided and therapy should be reassessed at every medical consultation, considering that the impact of the medication burden can be amplified.

It is important to distinguish between numerical polypharmacy and inappropriate medication use. Several studies have shown that the use of five or more medications may be clinically appropriate in older adults with multiple chronic conditions, particularly when treatments are evidence-based [6,22]. Nevertheless, even appropriate polypharmacy may increase vulnerability to adverse outcomes due to cumulative drug effects, treatment burden, and reduced physiological reserve [11,21,23]. Our findings therefore should not be interpreted as implying that all polypharmacy is inappropriate, but rather that higher medication burden identifies a subgroup of older adults with increased clinical vulnerability.

A significant strength of this study is the utilization of a substantial sample of older adults derived from the SABE Colombia survey. While the cohort specifically represents a major geographic region characterized by a high prevalence of aging, its size and depth offer robust epidemiological insights that transcend the limitations of small-scale or localized studies. Furthermore, the longitudinal design, encompassing a follow-up period of over six years, enabled the identification of clear temporal sequences between baseline medication burden and subsequent mortality. By demonstrating that the exposure preceded the outcome, this study provides a more rigorous assessment of these associations than traditional cross-sectional investigations, although the observational nature of the data precludes direct causal inferences.

The study’s methodological framework follows established epidemiological standards for longitudinal survival analysis. By employing Cox proportional hazards models adjusted for key confounders—such as age, sex, multimorbidity, and functional dependency—the analysis isolates the independent contribution of medication burden to mortality risk. While this statistical approach is consistent with prior research, the specific categorization into moderate polypharmacy (5–9 medications) and excessive polypharmacy (≥10 medications) offers a nuanced clinical perspective. These results suggest a non-linear relationship where the risk of mortality is markedly accentuated at higher thresholds of drug consumption. Consequently, this stratified analysis provides a pragmatic evidence base to support targeted medication review and deprescribing strategies in clinical practice.

This study has several limitations. First, polypharmacy was assessed at a single time point (baseline). As we lacked data on the duration of treatment or changes in medication regimens during the follow-up period, we could not account for time-varying exposure. This may introduce non-differential misclassification, potentially underestimating the true association. Furthermore, while interviewers verified prescriptions and packaging to minimize recall bias, reliance on self-report for some participants remains a limitation. Second, we did not assess medication appropriateness using explicit criteria, which limits our ability to distinguish appropriate from inappropriate polypharmacy. Third, although the sample size was large, recruitment was restricted to four of thirty-two departments, which limits the generalizability of the findings to the national level.

Moreover, reliance on self-reported data introduces the possibility of recall bias, particularly with respect to medication use, which may have influenced the accuracy of polypharmacy exposure. In addition, the study did not stratify outcomes by type of comorbidity, precluding a more granular analysis of disease-specific risks. The complexity of the Colombian healthcare system, which comprises diverse insurance schemes, necessitated specific eligibility criteria for outcome adjudication. Participants enrolled in ‘special regimes’—including military personnel and certain government employees—were excluded from the final analysis, as their vital status is not recorded in the ADRES database. This exclusion accounted for 283 individuals, representing a marginal attrition rate of 6.8% of the initial sample. While these participants were omitted to ensure the integrity and verifiability of the mortality data, the remaining cohort provides a robust representation of the population covered by the primary contributory and subsidized health insurance regimes.

Additionally, although we adjusted for key health indicators such as multimorbidity and functional dependence which capture a significant portion of the variance related to overall health status, specific geriatric domains such as cognitive status, depression, nutritional risk, and healthcare utilization frequency were not included in the final adjustment. Therefore, the possibility of residual confounding cannot be ruled out.

Finally, the interpretation of these findings necessitates a nuanced consideration of potential indication bias and reverse causation. While polypharmacy is a documented risk factor, it also functions as a surrogate marker for clinical complexity, frailty, and underlying disease severity, all of which independently contribute to mortality risk. Consequently, individuals with the most precarious health status are inherently predisposed to both higher medication counts and an increased baseline probability of death. Although our analysis rigorously adjusted for multimorbidity and functional status—two primary drivers of pharmacological accumulation—the possibility of residual confounding from unmeasured clinical variables cannot be entirely dismissed. Therefore, these results should be interpreted as robust longitudinal associations that characterize the risk profile of older adults with high medication burdens, rather than as a direct causal pathway.

## Conclusions

This longitudinal study demonstrates that polypharmacy is a robust and independent predictor of all-cause mortality among community-dwelling older adults in Colombia. Our findings reveal a non-linear risk, where the association with mortality is significantly accentuated in cases of excessive polypharmacy (≥10 medications), even after rigorous adjustment for multimorbidity and functional status. These results underscore that medication burden acts as a critical marker of clinical vulnerability in aging populations within middle-income settings. Consequently, there is an urgent need to transition from purely numerical medication counts to structured clinical reviews. Implementing proactive deprescribing strategies and optimizing therapeutic regimens are essential steps to mitigate the adverse consequences of pharmacological accumulation and to promote safer prescribing practices in resource-constrained healthcare systems.

## List of abbreviations

ADRES: Administrator of Resources of the General Social Security System in Health
B-ADL: Basic Activities of Daily Living
CI: Confidence Interval
COLCIENCIAS: Ministry of Health and Social Protection and the Administrative Department of Science, Technology, and Innovation
HR: Hazard Ratio
I-ADL: Instrumental Activities of Daily Living
IQR: Interquartile Range.
No: Number
Polypharmacy_5: use of five or more medications
Polypharmacy_10: use of ten or more medications.
SABE: Survey on Health, Well-Being, and Aging
SD: Standard Deviation
VIF: Variance Inflation Factor

## Acknowledgements

The study expresses its sincerest gratitude to all those who participated in the questionnaire.

## Authors’ contributions

H.D.G: Study conception and design, statistical analyses, drafting of the manuscript. C.G.B: Study conception and design, statistical analyses, drafting of the manuscript. J.H.D: Contribution to study design, data interpretation, and critical revision of the manuscript.

P.H: Contribution to study design. C.V: Contribution to study design.

L.A.T: Data analysis, interpretation, and manuscript revision.

C.L.C: Supervision of the study, data interpretation, and critical revision of the manuscript.

All authors read and approved the final version of the manuscript.

## Data availability

The data that support the findings of this study are available on request from the corresponding author. The data are not publicly available due to privacy or ethical restrictions.

## Declarations

### Ethical approval and consent to participate

The study was conducted in accordance with the principles of the Declaration of Helsinki (2013) for research involving human participants [24] and received approval from the Ethics Committee of the Universidad Pontificia Bolivariana on October 4, 2021. Data use complied with the guidelines of the Colombian Ministry of Health and Social Protection (MSPS), which grants end users access to the results of surveys developed within the framework of the National System of Population Studies and Health Surveys. All microdata were anonymized to safeguard the confidentiality of observational units, ensuring that no individual-level information could be inferred. The use of these data is permitted provided that appropriate acknowledgment is given to the MSPS and that researchers from participating institutions are recognized as co-authors for their active contribution to the survey. Consequently, individual informed consent was not required for this study, as it had been obtained at the time of survey implementation.

### Consent for publication

Not applicable.

